# Current infectious disease research practices with forcibly displaced people in the top ten low- and middle-income host countries: A Systematic Review

**DOI:** 10.1101/2024.09.24.24314319

**Authors:** Neila Gross, Maia C. Tarnas, Rashmina J. Sayeeda, Carly Ching, David Flynn, Muhammad H Zaman

## Abstract

Infectious disease research is essential for disease prevention and management within refugee camps and informal settlements. We aim to assess the state of infectious disease research with displaced communities in the top ten refugee-hosting low- and middle-income countries. We searched three journal databases for primary research that explicitly included refugees or was conducted in a refugee camp, informal settlement, or displaced people-serving hospital and focused directly on an infectious disease following PRISMA guidelines. Forty studies (out of 1,179) met the inclusion criteria. Common research challenges included population mobility, limited external validity, and low recruitment. No studies included the community in the initial study conception or investigated the research impact on the community. Community involvement was often through community health workers (45%). Of the 18 studies that studied a resource-based intervention, 20% explicitly noted that the intervention was unsustainable. Such context-specific considerations are vital in research with displaced communities.

## INTRODUCTION

The United Nations Refugee Agency (UNHCR) estimates that in 2022, 108.4 million people remained forcibly displaced. This included both refugees (people who are housed outside their country of citizenship) and internally displaced persons (communities that are displaced within their own countries). Of the 29.4 million documented as refugees, 76% are hosted in low- and middle-income countries (LMICs).^1^ The number of displaced individuals is expected to grow due to increasing poverty, insecurity, and declining access to essential services due to conflict and climate change-related environmental disasters.^2^ Displaced individuals are more susceptible to infectious diseases, often stemming from poor housing, insufficient access to healthcare and water, sanitation, and hygiene (WASH) infrastructure, environmental exposures, and overcrowding.^3^ Given this, research on infectious disease prevention, management, and control within the context of refugee camps and informal settlements is essential for enhancing public health preparedness and response. However, it is critical that such research does not further increase the harm or vulnerability of groups that already face compounding marginalization.^4^

Several organizations, including WHO and the Refugee Studies Center, have outlined frameworks for conducting ethical research with refugees or displaced persons.^5,6^ Most emphasize the involvement of displaced people throughout the research process and the importance of implementation research with long-term programming. Other humanitarian organizations that operate frequently in refugee camps, such as UNICEF and Médecins Sans Frontières, have broad ethical research guidelines that are not specific to working with displaced communities.^7,8^ Though there is apparent agreement in the overarching framing of how research involving refugees and other types of displaced people *should* be conducted, the extent to which this is actually done is unclear.^9,10^ Notably lacking is how to involve the community before the research agenda is set, how to iteratively (and collaboratively) develop the research topic and design, and logistical guidance on working with mobile populations.

These ethical considerations are especially critical in LMICs, where the health sciences have a notorious history of extractive research.^11–13^ In host countries that neighbor refugees’ country of origin, there may be increased scrutiny of refugees because of ongoing conflict, sociopolitical tensions, and xenophobia;^10^ infectious disease research must be aware of its potential to widen such divides. In this systematic review, we aim to assess the current state of research done to prevent, treat, and manage infectious diseases within refugee camps and informal settlements in the ten LMICs that host refugees and other types of forcibly displaced people.^14,15^ We emphasize how research has been conducted and the extent to which the community was involved throughout the process.

## METHODS

We conducted a systematic review of infectious disease prevention, treatment, and control research in refugee camps and informal settlements in the top ten refugee-hosting LMICs in 2023, as defined by the UNHCR.^14,15^ These countries are: 1. Iran (3.4 million refugees); 2. Pakistan (2.1 million); 3. Uganda (1.5 million); 4. Bangladesh (1 million); 5. Sudan (900,000); 6. Ethiopia (900,000); 7. Lebanon (800,000); 8. DRC (500,000); 9. Kenya (500,000); and 10. Cameroon (500,000). This review follows PRISMA guidelines and is registered on PROSPERO (CRD42023461567).

### Search Strategy and Inclusion Criteria

We searched PubMed, Embase, and Web of Science using MeSH terms (complete search strategy available in the **appendix**) on August 4th, 2023. A full protocol can be found on the PROSPERO website or in the appendix. We did not use a date or language filter in our initial search. To be included, studies had to be primary research, explicitly include refugees or be conducted in a refugee camp, informal settlement, or displaced people-serving hospital, and focus directly on at least one infectious disease. We included informal settlements because an estimated 88% of refugees live outside of camps, often in substandard urban dwellings.^16^ Papers that focused on topics that may have a downstream, indirect, or implied effect on infectious diseases were excluded. After deduplication, three authors (NG, MCT, and RJS) conducted a blind title and abstract screening followed by a full-text review using the web-based application Rayyan.^17^

### Data Extraction and Analysis

Following the full-text review, we extracted data on study location, participants, purpose and design, data collection, ethical considerations, intervention (if appropriate), and any other relevant information into a standardized spreadsheet. We specifically assessed how researchers involved the study population during the study conception and design. To measure intervention sustainability, we extracted information on explicit discussions of intervention longevity, cost-effectiveness, and appropriateness of intervention for the context. To avoid reporting bias, two authors extracted information independently (NG and MCT) and relayed any discrepancies following the extraction.

### Bias Assessments

We assessed studies’ risk of bias using the Mixed Methods Appraisal Tool (MMAT).^18^ For each study design type, the MMAT poses five criteria to assess bias risk. Studies that met at least four out of five criteria were considered high quality (i.e., low risk of bias).^19^ Two authors (MCT and NG) independently completed this analysis, and discrepancies were resolved by comparing textual evidence.

We also assessed the studies for publication bias. Outcomes were defined as positive, negative, and mixed (including both positive and negative results). Medical interventions were considered positive if there was a statistical difference between the intervention and placebo groups. Survey studies had a positive outcome if researchers were able to properly administer and complete the survey and found significant correlations resulting from the survey.

## RESULTS

### Screening Results

Our initial searches returned 1,179 total results (**Figure 1**). Of the 1,036 that remained following deduplication, 976 were removed following title and abstract review. Twenty papers were excluded after full-text screening. Based on this, 40 papers met the review’s inclusion criteria (**Table 1**).

**Figure 1:**
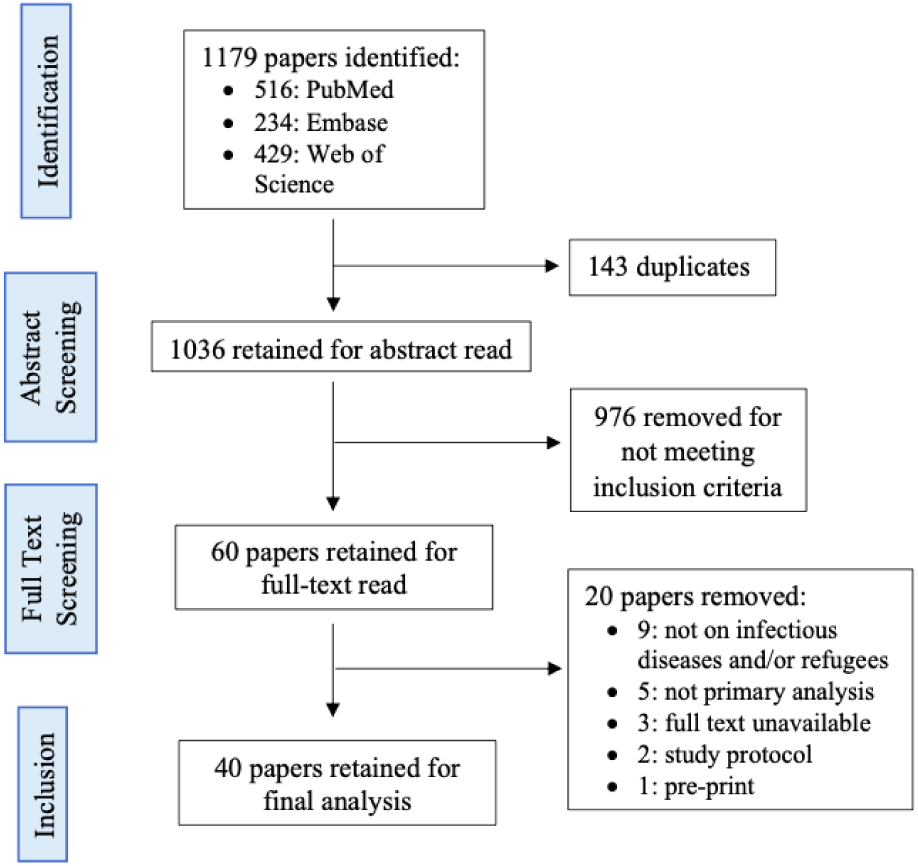
Study selection flowchart.

**Table 1:** Details of studies included in this review.

| Citation | Location | Disease focus | Study design | Topic of study/intervention |
| --- | --- | --- | --- | --- |
| Mekonnen et al.<sup>20</sup> | Ethiopia | Diarrhea | RCT | Hygiene promotion to reduce childhood diarrhea |
| Rego et al.<sup>21</sup> | Bangladesh | Diarrhea | Cross-sectional | Comparing diarrhea measurement tools |
| Logie et al.<sup>22</sup> | Uganda | COVID-19 | Cross-sectional | COVID-19 vaccination acceptability |
| Sanders et al.<sup>23</sup> | Sudan | Trachoma | Cross-sectional | Measuring trachoma prevalence |
| Zohura et al.<sup>24</sup> | Bangladesh | Diarrhea (cholera) | Mixed methods | Development of a cholera response program |
| Ali et al.<sup>25</sup> | Bangladesh | Diarrhea (cholera) | RCT | Cholera vaccine effectiveness |
| Mohiuddin Chowdhury et al.<sup>26</sup> | Bangladesh | COVID-19 | RCT | Clinical recovery from COVID-19 |
| Brooks et al.<sup>27</sup> | Bangladesh | Pneumonia and diarrhea | RCT | Zinc supplementation to reduce disease incidence |
| Brown et al.<sup>28</sup> | Pakistan | Pneumonia | Case-control | Associations with environmental factors |
| Mohiuddin Chowdhury et al.<sup>29</sup> | Bangladesh | COVID-19 | RCT | Treatment of severe COVID-19 |
| Faruque et al.<sup>30</sup> | Bangladesh | Diarrhea | Cross-sectional | Characteristic of patients hospitalized with diarrhea |
| Wolday et al.<sup>31</sup> | DRC | Malaria | Cross-sectional | Measuring the resistance of *P. falciparum* to antimalarials |
| van der Kop et al.<sup>32</sup> | Kenya | HIV | RCT | Patient retention in HIV care |
| Stein et al.<sup>33</sup> | Uganda | COVID-19 | Mixed methods | Impact of one-time cash transfer |
| O’Laughlin et al.<sup>34</sup> | Uganda | HIV | Cross-sectional | Associations with HIV diagnosis |
| Rowland et al.<sup>35</sup> | Pakistan | Malaria | Randomized crossover trial | Insecticide treatment of cattle |
| Rowland et al.<sup>36</sup> | Pakistan | Malaria | RCT | Insecticide-treated chaddars and top-sheets |
| Graham et al.<sup>37</sup> | Pakistan | Malaria | RCT | Comparison of insecticides for top-sheet treatment |
| Howard et al.<sup>38</sup> | Pakistan | Malaria | RCT | Malaria treatment |
| Kaul et al.<sup>39</sup> | Kenya | HIV / STI | RCT | Antibiotic prophylaxis for STIs and HIV |
| Kimani et al.<sup>40</sup> | Kenya | Malaria | RCT | Insecticide-treated clothes |
| Kolaczinski et al.<sup>41</sup> | Pakistan | Malaria and CL | Case study | Shift to subsidized sale of bed nets |
| Kolaczinski et al.<sup>42</sup> | Pakistan | Malaria | RCT | Malaria treatment |
| Larson et al.<sup>43</sup> | Bangladesh | Diarrhea | RCT | Zinc supplementation after diarrheal treatment |
| Leslie et al.<sup>44</sup> | Pakistan | Malaria | RCT | Compliance with malaria treatment |
| Luby et al.<sup>45</sup> | Pakistan | Diarrhea, impetigo, ARI | RCT | Handwashing promotion with soap |
| Luby et al.<sup>46</sup> | Pakistan | Diarrhea | RCT | Handwashing promotion with soap |
| Luoto et al.<sup>47</sup> | Bangladesh | Diarrhea | RCT | Water treatment products and education |
| Murphy et al.<sup>48</sup> | Pakistan | Diarrhea | Non-randomized controlled trial | Trial of wheat-based oral rehydration solution |
| Ngure et al.<sup>49</sup> | Kenya | HIV | RCT | 6-month PrEP dispensing with self-HIV testing |
| Qadri et al.<sup>50</sup> | Bangladesh | Diarrhea | RCT | Vaccine safety and immunogenicity |
| Rahman et al.<sup>51</sup> | Bangladesh | Diarrhea and ARI | RCT | Zinc and vitamin A supplementation |
| Rowland et al.<sup>52</sup> | Pakistan | Malaria | RCT | Pyrethroid-impregnated bed nets |
| Rowland et al.<sup>53</sup> | Pakistan | Malaria | RCT | DEET mosquito repellent soap |
| Rowland and Durrani<sup>54</sup> | Pakistan | Malaria | RCT | Malaria treatment |
| Mitra et al.<sup>55</sup> | Bangladesh | Diarrhea, dysentery, ARI | RCT | Long-term oral iron supplementation |
| Najnin et al.<sup>56</sup> | Bangladesh | ARI | RCT | Handwashing intervention |
| Owais et al.<sup>57</sup> | Pakistan | DTP/Hep B vaccine | RCT | Low-literacy educational intervention |
| Pickering et al.<sup>58</sup> | Bangladesh | Diarrhea | RCT | Water treatment methods |
| Pickering et al.<sup>59</sup> | Bangladesh | Diarrhea | RCT | In-line drinking water chlorination |

RCT: randomized controlled trial; HIV: human immunodeficiency virus; STI: sexually transmitted infection; CL: cutaneous leishmaniasis; ARI: acute respiratory infection; PrEP: pre-exposure prophylaxis

### Study Characteristics

The included studies were published between 1995 and 2023, and most were located in Bangladesh (n=15)^21,24–27,29,30,43,47,50,51,55,56,58,59^ and Pakistan (n=15)^28,35–38,41,42,44–46,48,52–54,57^ (**Table 2**). No studies were conducted in Lebanon, Iran, or Cameroon. Sixteen studies20,^21,24,25,27,30,43,45–48,50,51,55,58,59^ (40%) focused on diarrheal diseases and 11^31,35–38,41,42,44,52–54^ (28%) on malaria. Most studies researched an intervention (85%, n=34),^20,24–27,29,31–33,35–59^ and most (70%, n=28)^20,25–27,29,32,36–40,42–47,49–59^ were randomized controlled trials (RCTs).

**Table 2:** Table of study location, year, and disease of focus.

|  | Diarrhea | Malaria | HIV/STD | COVID-19 | Acute resp. infection | Pneumonia | Other | Total |
| --- | --- | --- | --- | --- | --- | --- | --- | --- |
| **Bangladesh** | **12** | **0** | **0** | **2** | **3** | **1** | **0** | **18** |
| 1995-2000 | 1 | - | - | - | 1 | 1 | - | 2 |
| 2001-2006 | 3 | - | - | - | 1 | - | - | 5 |
| 2007-2012 | 2 | - | - | - | - | - | - | 2 |
| 2013-2018 | 1 | - | - | - | - | - | - | 1 |
| 2019-2023 | 5 | - | - | 2 | 1 | - | - | 9 |
| **Pakistan** | **3** | **10** | **0** | **0** | **1** | **1** | **3** | **18** |
| 1995-2000 | 1 | 3 | - | - | - | - | - | 4 |
| 2001-2006 | 2 | 5 | - | - | 1 | - | 2* | 10 |
| 2007-2012 | - | 2 | - | - | - | - | 1** | 3 |
| 2013-2018 | - | - | - | - | - | - | - | 0 |
| 2019-2023 | - | - | - | - | - | 1 | - | 1 |
| **Uganda** | **0** | **0** | **1** | **2** | **0** | **0** | **0** | **3** |
| 1995-2000 | - | - | - | - | - | - | - | 0 |
| 2001-2006 | - | - | - | - | - | - | - | 0 |
| 2007-2012 | - | - | - | - | - | - | - | 0 |
| 2013-2018 | - | - | 1 | - | - | - | - | 1 |
| 2019-2023 | - | - | - | 2 | - | - | - | 2 |
| **Kenya** | **0** | **0** | **4** | **0** | **0** | **0** | **0** | **4** |
| 1995-2000 | - | - | - | - | - | - | - | 0 |
| 2001-2006 | - | - | 2 | - | - | - | - | 2 |
| 2007-2012 | - | - | - | - | - | - | - | 0 |
| 2013-2018 | - | - | 1 | - | - | - | - | 1 |
| 2019-2023 | - | - | 1 | - | - | - | - | 1 |
| **DRC** | **0** | **1** | **0** | **0** | **0** | **0** | **0** | **1** |
| 1995-2000 | - | 1 | - | - | - | - | - | 1 |
| 2001-2006 | - | - | - | - | - | - | - | 0 |
| 2007-2012 | - | - | - | - | - | - | - | 0 |
| 2013-2018 | - | - | - | - | - | - | - | 0 |
| 2019-2023 | - | - | - | - | - | - | - | 0 |
| **Sudan** | **0** | **0** | **0** | **0** | **0** | **0** | **1** | **1** |
| 1995-2000 | - | - | - | - | - | - | - | 0 |
| 2001-2006 | - | - | - | - | - | - | - | 0 |
| 2007-2012 | - | - | - | - | - | - | - | 0 |
| 2013-2018 | - | - | - | - | - | - | - | 0 |
| 2019-2023 | - | - | - | - | - | - | 1*** | 1 |
| **Ethiopia** | **1** | **0** | **0** | **0** | **0** | **0** | **0** | **1** |
| 1995-2000 | - | - | - | - | - | - | - | 0 |
| 2001-2006 | - | - | - | - | - | - | - | 0 |
| 2007-2012 | - | - | - | - | - | - | - | 0 |
| 2013-2018 | - | - | - | - | - | - | - | 0 |
| 2019-2023 | 1 | - | - | - | - | - | - | 1 |
| **Total** | **17** | **13** | **5** | **4** | **4** | **2** | **4** |  |

\*: impetigo and leishmaniasis, \*\*: diphtheria, tetanus, pertussis, and hepatitis B, \*\*\*: trachoma.

### Challenges in conducting research in displaced person settings

Researchers reported several challenges in conducting research in these settings (**Figure 2**). Eleven studies (27.5%) cited high population mobility as a challenge,^21,23,25,39,40,42,45,47,55,56,59^ with five studies (12.5%) requiring that study participants remain in the camp throughout the study’s duration.^21,39,40,42,45^ One study reported that 67% of the baseline population either migrated or died throughout the duration of the study.^25^ Limited external validity (15%, n=6),^22,25,30,34,43,59^ low recruitment (12.5%, n=5),^26,29,31,42,58^ and attrition not specific to population mobility (10%, n=4)^33,49,51,58^ were also challenges. Several studies were either interrupted or ended prematurely due to civil unrest,^21^ natural disasters,^21^ COVID-19,^24,33^ resource constraints,^42,59^ or operational constraints.^33^

**Figure 2:**
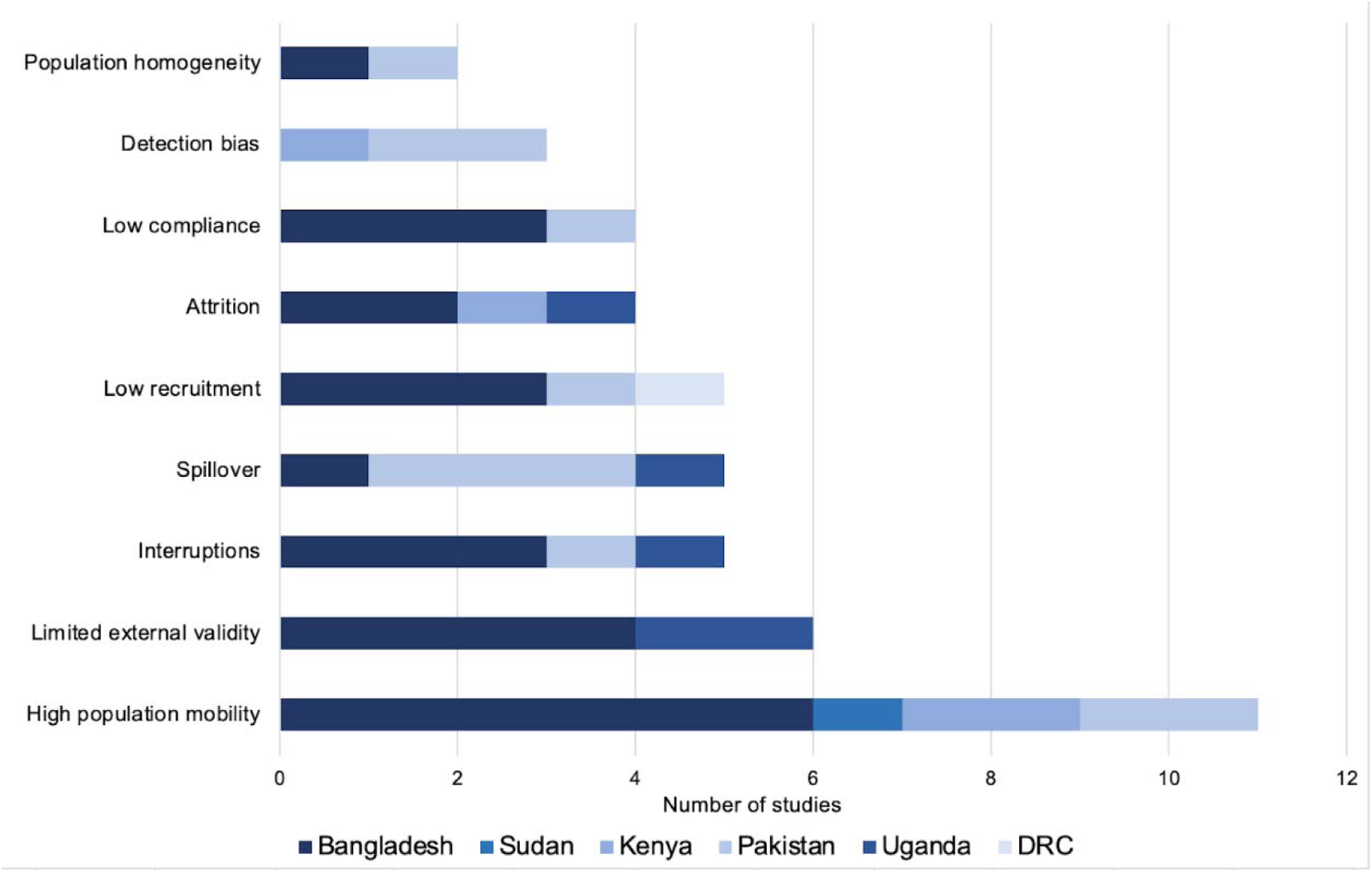
Research challenges in refugee camps and informal settlements by country of study. Studies may have mentioned multiple challenges.

Camp organization posed several difficulties. The density of the setting led to spillover concerns in five studies (12.5%).^25,33,44,48,57^ One study, however, found that the camp’s structured nature aided in the study’s survey distribution.^23^ Another study was unable to easily locate households because the official camp map (from the UNHCR) was outdated.^33^ Concerns that the relative homogeneity (specifically, socioeconomic homogeneity) of the population blunted observed associations were expressed in two studies.^28,43^ The authors of one study noted that the complex household composition common in their study setting made proper implementation of the intervention difficult.^33^

In studies that tested an intervention, four (10%) noted low compliance or uptake as a challenge.^27,36,47,56^ One study credited the low compliance to cultural norms, as the study called for participants not to wash *chaddars* (a cloth used for head covering) that had been treated with insecticide despite norms calling for regular washes.^36^

Detection bias was a significant concern noted in several studies.^39,46,48^ This arose over the belief that an intervention’s observed success may not be due to the intervention itself but rather from the increase in overall baseline care, interactions with study staff, regular house visits, and provision of material goods. Lastly, other challenges of note included language barriers^23,33^ and lack of health or camp records.^23,59^

### Ethical Considerations and Community Involvement

Of the 33 studies (83%) that received ethical review board approval, 32 received approval from an ethical review board in the country where the research was performed. Three studies had independent data and safety boards with which researchers met throughout the trial.^32,43,50^ Only two studies either compensated participants^39^ or were cognisant of the potential monetary burden of participating.^42^ Lastly, 26 studies (65%) did not give a specific reason for conducting the research in a refugee camp or with displaced people.^20–22,24,26,27,29,31,32,35,42,43,45–51,53–59^

No studies mentioned including the community in the study conception or initial design process. Eighteen studies (45%) directly involved the community during the study (beyond using local health centers);^20,23,24,32,33,35,36,39–41,44–46,52,55–57,59^ this was most commonly done via community health workers (CHWs) (25%, n=10)^32,36,40,41,44,45,52,55–57^ or field workers hired from the community (7.5%, n=3).^33,35,46^ One study used a translator from the participating camp,^23^ another used peer educators,^39^ and a third used local environmental health workers.^20^ Community leaders were involved in three studies by providing informed consent^45,46^ and assisting in community mobilization.^40^ The community was directly involved in implementing the intervention in two studies; in both studies, participants treated items with insecticide under the study team’s supervision.^35,36^

The community was used to inform intervention components in three studies. Importantly, in none of these studies was the community involved at the onset, but rather once the initial conceptualization had already been completed. In one study, the researchers conducted semi-structured interviews with community members to inform intervention modifications.^24^ Similarly, another study used questionnaire results to inform intervention pricing.^41^ The community was involved a second time in lowering the price further. In the third study, local ethics experts were consulted to determine an appropriate control.^59^ However, it is unclear whether ‘local’ refers to community members or if it was being used more broadly.

### Intervention Feasibility and Sustainability

Forty-five percent (n=18) of intervention-based studies provided resources (beyond medical care and/or medication) to participating individuals, households, or communities (**Figure 3**).^20,24,33,35–37,39–41,45–47,52,53,56–59^ This included educational materials (20%, n=8),^24,33,45–47,53,57,59^ bed nets, insecticide, or other mosquito repellents (17.5%, n=7),^35–37,40,41,52,53^ water treatment and storage supplies (12.5%, n=5),^24,47,56,58,59^ hand washing materials (10%, n=4),^20,24,45,56^ a cash transfer,^33^ and condoms.^39^ In all but one of these studies, the materials were provided to participants for free and were restocked throughout the study. The one study that did charge for the intervention (insecticide-treated bednets) was purposefully doing so to assess the feasibility of transitioning away from free net distribution and the study planned for free annual retreatment.^41^ Outside of this, no other studies discussed plans for sustained provision of the intervention(s), even when they were successful. In fact, several studies (20%, n=8) explicitly noted that the tested interventions (including one clinical intervention) were not feasible in the studied location.^20,24,39,45,46,49,58,59^ There were several reasons that studies considered their interventions unfeasible, but primarily included unavailability of resources and cost (**Figure 3**).

**Figure 3:**
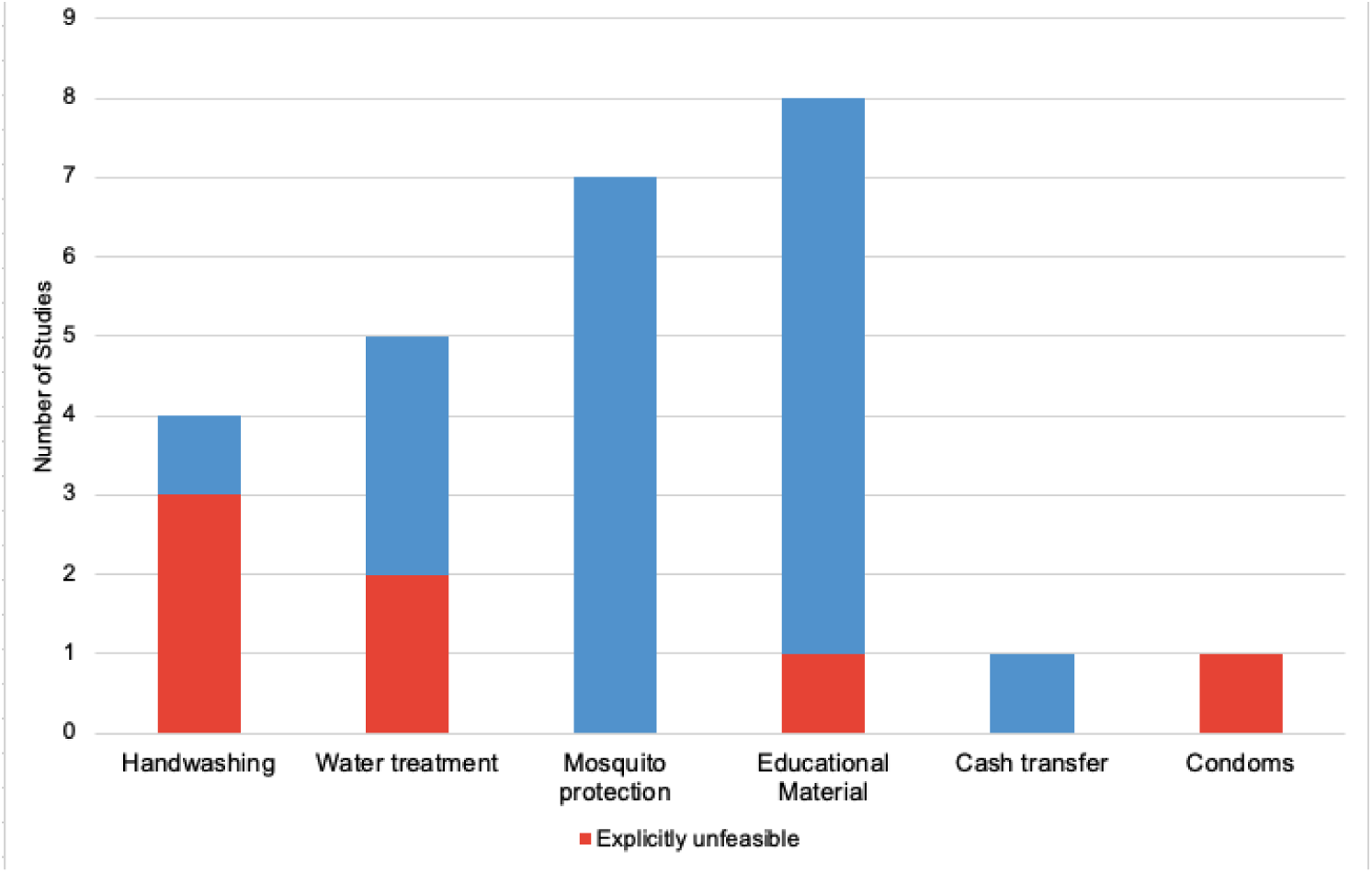
Frequency of intervention types in relevant intervention-based studies included in the review, and the proportion of those studies explicitly considered their intervention unfeasible. Individual studies may have had multiple interventions.

In two studies, the materials used for the intervention (a tablet for water treatment and an insecticide) were not available within the studied country.^40,58^ The tested automated system in the first study was too expensive to be successfully scaled up and affordable in that location.^58^ Similarly, two handwashing interventions were successful within the studies but were both described as “prohibitively expensive” for large-scale implementation.^45,46^ Another study tested a passive water treatment system that required water points to be connected to water storage tanks; only 25% of water points in the studied area met this standard.^59^ Two studies noted concerns over whether the successful intervention material would be available once the study team stopped providing it.^20,24^ In one of these studies, the research team noted that the intervention’s effectiveness fell three months after the study ended, perhaps due to “limitations in infrastructure or resources.”^20^ Another study provided “state-of-the-art” health services that were “well above the standard of care for [the] region” and were likely unable to be replicated following the study’s conclusion.^39^

Ten studies (25%) tested pharmacological or clinical interventions.^27,38,42,44,48–51,54,55^ In three studies (7.5%), researchers tested the efficacy of official (at the time) malaria treatment plans,^38,42,54^ two of which had been implemented despite no supporting evidence.^38,54^ The policies were all ineffective; one study’s findings were used to significantly alter official malaria treatment guidance.^42^ Two other studies had the potential for policy implications regarding medication provision, though no related policy changes were discussed in the articles.^44,49^

Two interventions had the potential to be cost-effective but were not at the time of study, given large-scale implementation and development issues.^57,58^ Conversely, several (17.5%, n=7) studies tested interventions that were already considered cost-effective.^27,35,36,40,49,52,53^ In four of these studies, the intervention was tested explicitly because a cost-effective measure was needed to replace the status quo.^35,36,40,52^ Two studies considered cultural norms when choosing an intervention.^36,48^ In one, researchers tested a homemade wheat-based oral rehydration solution because it allowed women to remain in the home (as is the norm), thereby making it a more accessible solution than typical oral rehydration salts.^48^ Based on this study, this homemade solution became an approved treatment in the study district.^48^

### Bias Assessments

Across the five criteria for RCTs in the MMAT, 14 of the 29 RCTs (including a crossover RCT) were considered high quality.^20,25,27,32,38,39,42,43,49–51,55,57,59^ Both quantitative non-randomized studies were high quality,^28,48^ as were both mixed methods studies^24,33^ and the single qualitative study.^41^ Lastly, four of the six quantitative descriptive studies were considered high quality. The full risk of bias assessment can be found in the **appendix**.^21–23,30^

For publication bias, two studies (5%) had mixed results depending on the age of the participant,^25,36^ twenty-three studies (57%) reported positive outcomes for their interventions,^20,23,24,27,28,30,31,34,35,37,40–43,45,46,48–52,57,59^ and 15 (37.5%) reported negative outcomes.^21,22,26,29,32,33,38,39,44,47,53–56,58^ Among the 40 included studies, one individual (M. Rowland) was either the first or senior author on ten studies.^35–38,41,42,44,52–54^ Another author (S. Luby) was either the first or senior author on five studies, meaning that two authors were responsible for nearly 40% of the studies included in this review.^45,46,56,58,59^

## DISCUSSION

To our knowledge, this review is the first to assess the landscape of infectious disease research conducted in refugee camps and informal settlements in LMICs. Despite existing guidelines for conducting research with refugees or other types of displaced people that emphasize community participation and intervention sustainability, we found that most studies fell short across several metrics. None of the included studies involved the participating community in the study conception or design process. Figure 3 illustrates that in 20% of studies, researchers tested interventions that they explicitly noted were not feasible to implement in the studied context. Such findings are stark reminders that while there may be consensus on what ethical research should look like in these settings, the implementation of such guidelines is frequently lacking.

Several studies noted particular difficulties with conducting research in refugee camps and informal settlements, many of which are likely inherent to these settings, such as high population mobility. Such realities are perhaps foreseeable in these settings, though there was little discussion of how the research was adapted to fit them. Instead, many studies required that participants alter their behaviors for the study: for example, several studies required that participants stay in the camp throughout the duration. Such requirements highlight tensions between the ostensible impermanence of camp settings, unique population dynamics, and rigid research methodologies that must be addressed both explicitly and under ethical guidance. Similarly, research within a camp (and especially that which involves an intervention) inherently changes camp dynamics through the introduction of resources, attention, and funding. However, only three studies noted concerns over detection bias, despite many of the intervention-based studies providing resources that may otherwise be difficult to obtain for the community. Overwhelmingly, studies in this review lacked a critical discussion of how the research’s end may impact the community, especially when the conclusion of the research corresponds to a concurrent conclusion in resource availability.

There was also a noticeable imbalance between countries in which research was conducted. Despite Iran hosting the most refugees for an LMIC, there were no Iran-based studies that met our inclusion criteria. Pakistan and Bangladesh were overrepresented in the included studies. Notably, most of the studies in Bangladesh involved the International Center for Diarrheal Disease Research, Bangladesh (ICDDR, B), indicating that there is strong institutional support for related research in Bangladesh. This may also explain the heavy focus on diarrheal disease in the Bangladesh-based research.

Less than half of the studies in this review involved the community during the study period, and the majority of this involvement was through CHWs. The benefits of using CHWs have been well-documented in the literature, and such inclusion is important.^60^ However, there were only three studies that explicitly discussed researchers being informed *by* the community. Such iterative collaboration on the research process is essential for the co-construction of research and likewise may aid in the development of projects that meet the community’s highest perceived needs.

This review has several limitations. It is possible that we missed studies that meet the inclusion criteria due to different or changing terminology. We attempted to mitigate this through the use of comprehensive search terms and checking these terms with relevant experts. It is also possible that we misclassified some studies because the authors did not explicitly state the information in the text. For example, a study may have included community members in their research but did not explicitly state this in the final article, and we, therefore, did not count it as involving the community. We limited our data extraction to explicit mentions, however, to avoid subjective assumptions by our research team. We also viewed each of these studies in isolation, which may mask researchers’ prolonged involvement with certain communities. The articles in this review span nearly 30 years, during which time ethical standards of research, and indeed research practice generally, have evolved. However, we applied the same rigorous ethical expectations across each study, as the lack of formal guidelines should not be used to excuse unethical research practices.

Our findings highlight a critical and an unmet need for practical guidelines on conducting ethically sound research with displaced communities, both in LMICs and elsewhere. Though other guidelines exist, there is a disconnect with implementing them in these complex settings; this review helps indicate where these gaps between framework ideals and practical implementation often reside. The forced migration experience is singular, and more research must be attuned to the specific needs affected communities face. Moreover, more work must be done by researchers to critically assess their research, namely to critique the necessity of the research (who does it benefit?), the appropriateness of the intervention (is this intervention accessible and sustainable in this context?), the frequency of collaboration with the community (has the community-led this work?), and the effects of the work’s presence after the project is completed (how will this change the community?). These deliberations should be prominent in the published work from the project and, indeed, should be expected by journal editors and readers.

## CONCLUSION

In conclusion, research done in refugee camps and informal settlements in LMICs frequently lacks comprehensive ethical considerations (or relevant discussions of them) that are unique to working with these communities. Maintaining ethical standards is particularly crucial with displaced populations, where power imbalances and unique challenges exist. However, this makes their presence even more important, especially given the external challenges and marginalization faced by displaced communities. Reestablishing the health and well-being of participating displaced communities as the core priority of health research can be aided by more conscious efforts to conduct ethical research that is informed by - and adjusted to - the community itself.

## Data Availability

All data used for this study has been included in the manuscript or supplementary material.

## Acknowledgments

The authors would like to acknowledge Boston University’s Center for Forced Displacement.

## Author’s Contributions

NG: Investigation, Methodology, Formal Analysis, Writing-original draft, Writing-review and editing

MCT: Investigation, Methodology, Formal Analysis, Writing-original draft, Writing-review and editing

RS: Investigation, Methodology, Writing-review and editing

CC: Conceptualization, Writing-review and editing

DF: Methodology

MHZ: Conceptualization, Writing-review and editing

## DATA SHARING

All data used for this study has been included in the manuscript or supplementary material.

